# Impact of exercise training on sarcopenia associated with non-alcoholic fatty liver disease (NAFLD) in humans: A systematic review and meta-analysis

**DOI:** 10.1101/2020.09.05.20189100

**Authors:** Andrea Gonzalez, Mayalen Valero-Breton, Camila Huerta-Salgado, Oscar Achiardi, Felipe Simon, Claudio Cabello-Verrugio

**Author notes:** Corresponding author: Claudio Cabello-Verrugio. Laboratory of Muscle Pathology, Fragility, and Aging. Department of Biological Sciences. Faculty of Life Sciences. Universidad Andres Bello. Santiago, 8370146, Chile;.

## Abstract

**Objective:** To conduct a systematic review and meta-analyses to assess the efficacy of physical exercise on strength, muscle mass and physical function in adult patients with non-alcoholic fatty liver disease (NAFLD).

**Design:** We conducted a systematic review and meta-analysis of seven studies to investigate the effect of exercise training interventions in muscle strength, muscle mass and physical performance.

**Data sources:** We identified relevant randomised controlled trials (RCT) in electronic databases (PubMed, CINAHL and Scopus).

**Eligibility criteria:** We selected seven RCTs from 66 screened studies. The inclusion criteria were peer-reviewed and English writing articles that included adult patients with liver disease of non-alcoholic origin, applied resistance training, endurance training or both, and assayed at least one variable of sarcopenia.

**Results:** Physical performance criterion improved in the exercise groups (mean differences [MD] 8.26 mL/Kg*min [95% CI 5.27 to 11.24 mL/Kg*min], p < 0.0001) versus the control groups; muscle mass, determined as lean body mass (LBM), showed no evidence of the beneficial effects of exercise versus the control groups (MD 1.01 Kg [95% CI −1.78 to 3.80 Kg], p = 0.48); we did not include muscle strength, as none of the selected studies evaluated it.

**Summary/conclusion:** Exercise training is a useful intervention strategy to treat sarcopenia in patients with NAFLD; it increases their physical performance in the form of aerobic capacity but does not affect LBM. Future research should include muscle strength assessments and resistance training to evaluate the effects of exercise training on sarcopenia in NAFLD patients.

PROSPERO reference number CRD42020191471

## INTRODUCTION

Non-alcoholic fatty liver disease (NAFLD) is the most common type of chronic liver disease (CLD). It includes a broad spectrum of disorders ranging from accumulating lipids in the liver (steatosis) to the progressive inflammation denoted as non-alcoholic steatohepatitis (NASH) to advanced stages of damage such as fibrosis and cirrhosis.^1 2^

Sarcopenia is one of the most common complications associated with NAFLD, with a prevalence of 30—70%.^3^ According to the European Working Group on Sarcopenia in Older People (EWGSOP), sarcopenia diagnosis is based on three criteria: i) low muscle strength (the primary indicator of sarcopenia), ii) small muscle quantity or quality and iii) low physical performance.^4^ Sarcopenia negatively affects NAFLD progression;^5^ its severity is more pronounced in the advanced stages of NAFLD, considered as an independent predictor of pre- and post-liver transplant complications and mortality.^6 7^ Therefore, therapeutic interventions to prevent, revert or improve sarcopenia associated with NAFLD are essential.

Since sarcopenia involves a decline in muscle mass, strength and physical function, physical exercise is a promising tool for treating sarcopenia.^8^ Due to low-quality evidence, the results of studies that evaluate the effect of exercise training on patients with cirrhosis-associated sarcopenia are highly variable and inconclusive.^9^ Despite evidence that training has a positive effect on liver dysfunction parameters in patients with NAFLD,^10 11^ no studies evaluate the impact of training on sarcopenia parameters in NAFLD patients. In this context, a recent narrative review concluded that there is no consensus on the best type of exercise to improve muscle strength and physical function in NAFLD patients. Thus, more evidence is required to determine exercise specifications (type, intensity, frequency, supervised vs domiciliary).^2^

Considering all the antecedents, we conclude that the effects of exercise training on sarcopenia associated with NAFLD are poorly determined and that there is no consensus on the best type of exercise for these patients. Therefore, our objective was to conduct a systematic review and meta-analyses to assess the efficacy of physical exercise on strength, muscle mass and physical function in adult patients with NAFLD.

## METHODOLOGY

### Protocol and Registration

We conducted our systematic review and meta-analysis following the Preferred Reporting Items for Systematic Reviews and Meta-Analysis (PRISMA) statement. We registered the review protocol in the International Prospective Register of Systematic Reviews (PROSPERO reference number CRD42020191471), available from 3 September 2020.

### Search strategy

We performed a systematic search in PubMed, CINAHL and Scopus from March–May 2020. We included articles published from January 2000–March 2020 containing the critical concepts of sarcopenia, fatty liver disease, exercise and related words. The essential concepts of the search were different for each database. For example, for PubMed, we used the keywords (‘atrophic muscular disorders’ OR ‘muscle atrophy’ OR ‘muscle degeneration’ OR ‘muscle fiber atrophy’ OR ‘muscle fiber degeneration’ OR ‘muscle wasting’ OR ‘muscular wasting’ OR ‘muscular atrophy’ OR ‘muscular atrophies’ OR ‘muscular degeneration’ OR ‘sarcopenia’) AND (‘non-alcoholic steatosis’ OR ‘non-alcoholic fatty liver disease’ OR ‘fatty liver’ OR ‘hepatic fat’ OR ‘liver fibrosis’ OR ‘liver disease’ OR ‘fatty liver disease’ OR ‘obesity’) AND (‘exercise’ OR ‘physical activity’ OR ‘exercise intervention’ OR ‘training’) (see Supplementary Material 1). We filtered the results for clinical randomised controlled trials (RCTs). Then, AG reviewed all titles and abstracts from the articles selected and only downloaded those matching the inclusion criteria.

### Inclusion/exclusion criteria

The inclusion criteria are based on the study population, intervention, comparison question (control) and outcomes (PICO).^12^

- Investigate adults (18 or older) with liver disease of non-alcoholic origin (specifically, NAFLD or NASH) as confirmed by at least one of these parameters: hepatic biopsy, ultrasonography, computed tomography (CT), nuclear magnetic resonance spectroscopy (MRS), serum bile acids, gamma-glutamyl transpeptidase (GGT), aminotransferases, the ratio of aspartate transaminase (AST)/alanine transaminase (ALT) or bilirubin, alkaline phosphatase, dyslipidaemia)^13–17^ (**Population**).
- Apply resistance training, endurance training or both. Combined exercise is defined as interventions that simultaneously used resistance and endurance training (**Intervention**).
- Apply exercise alone or in combination with other interventions (educational, nutritional, etc.); must have a control group and a group with exercise only (to determine its independent effect) (**Control**). Control groups cannot have any intervention that could influence study outcome measures (e.g. nutritional or physical intervention). Moreover, participants of the exercise group must perform three or more sessions of training per week for at least four weeks to determine the chronic effect of exercise on the variables. Studies must conduct pre-and post-intervention assessments to identify changes in the variables.
- Assay at least one variable included in the consensus definition of sarcopenia:^2 18^ muscle strength, defined as the force generated through muscular contraction against an external load;^19^ muscle mass, defined as the part of total body mass composed of skeletal muscle tissue;^20^ and physical performance, defined as an objectively measured whole body function related with mobility.^21^ Thus, the studies must evaluate any of three criteria, as it was described in Table 1 (**Outcome**).
- Be a peer‐reviewed study
- Be written in English

Exclusion criteria:

- Bibliographic reviews, not controlled or randomised clinical trials, animal models
- The participants had comorbidities such as type II diabetes mellitus, viral hepatitis, cardiovascular disease, lung disease or neurological disease.
- Interventions that would interfere with identifying any exercise-mediated effects

Two researchers (CH-S and MV-B) independently reviewed the full-text version of the reports. They resolved discrepancies/disagreements by consensus – when this was not possible, two independent researchers acted as referees (AG and CC-V).

**Table 1.** Measurement of sarcopenia criteria in NAFLD patients: muscle strength, muscle mass and physical performance in clinical practice and research.

| Body composition<br>(muscular mass) | Muscular Strength | Physical Performance |
| --- | --- | --- |
| Extremity circumferences (Thigh, Arm) | Handgrip strength | 6-MWT (6-minute walk test) |
| Thigh US (ultrasound) | Knee flexion/extension | 2-MST (2-minute step test) |
| BIA (bioelectrical impedance analysis) | Dynamometer | CPET (cardiopulmonary exercise testing) |
| CSA (the cross-sectional area from magnetic resonance imaging) | 1 maximum repetition (1RM) | SPPB (Short physical performance Battery) |
| DXA (dual-energy x-ray absorptiometry) | 10 maximum repetition (10RM) | Usual gait speed |
| Anthropometry | Isokinetic evaluation | Chair stands |
| MAMA (middle-arm muscle area) | Peak expiratory flow (specific to respiratory) | Timed get-up-and-go test |
|  |  | Stair climb power test |

### Data extraction

We collected the data from seven final papers (n = 7), which we tabulated and ordered into a Microsoft Excel 2016 database. One researcher extracted the data, and two different researchers reviewed it to ensure data processing accuracy (Supplementary Material 2). The data included the following parameters: first author’s surname; publication year; number of participants by group; participants’ sex and age; method used to diagnose NAFLD and its severity; type, intensity, frequency and duration of the exercise intervention; sarcopenia criteria assessment methods. When studies presented intention-to-treat (ITT) values, we only extracted ITT data. Data extraction and synthesis for the outcome of interest are shown in the data synthesis section.

### TESTEX rating scale application

We assessed study quality using the TESTEX rating scale (Table 2).^22^ It includes criteria to assess methodological quality as well as ranking the whole article. The scale comprises 12 assessment criteria for a maximum score of 15 points. Higher ratings reflect better study quality and reporting^22^.

**Table 2.**
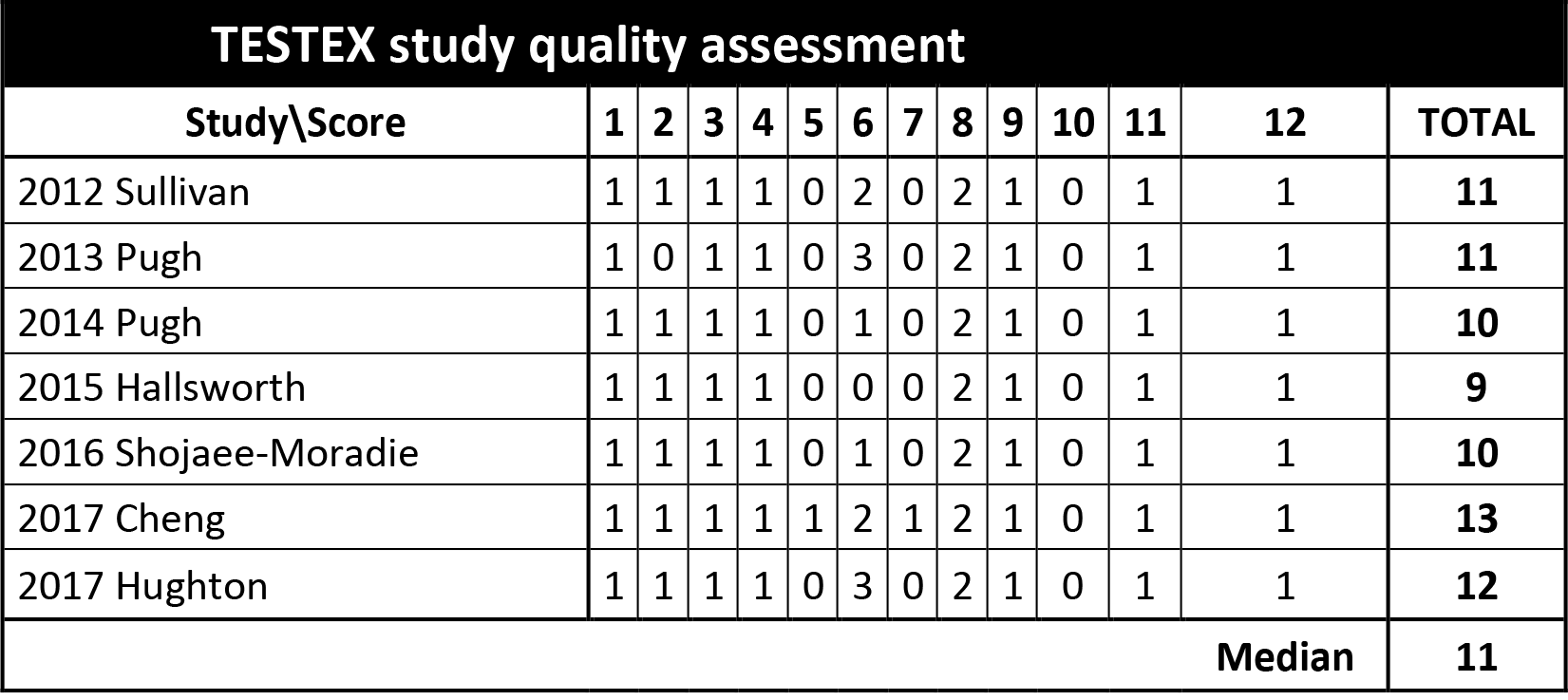
Study quality.

*Notes*. 1) eligibility criteria specified (1 point); 2) randomisation defined (1 point); 3) allocation concealment (1 point); 4) groups similar at baseline (1 point); 5) assessor blinding in study reporting (1 point); 6) outcome measures assessed in 85% of patients (3 points); 7) intention‐to‐treat analysis (1 point); 8) between‐group statistical comparisons reported (2 points); 9) point measures and measures of variability for all reported outcome measures (1 point); 10) activity monitoring in the control group (1 point); 11) relative exercise intensity remained constant (1 point); 12) exercise volume and energy expenditure (1 point).

TESTEX does not yet provide a validated cutoff score,^22^ so we categorised the studies depending on the median score.^10 23^ We classified studies above the median score as ‘high-quality’ and those below as ‘low-quality’. AG and OA performed TESTEX scoring separately. When different evaluations occurred, they discussed discrepancies until reaching a consensus score.

### Data synthesis

When at least three included studies reported the same outcome, we pooled data to perform a meta-analysis using Review Manager 5.4 (RevMan 5.4) as per the Cochrane Manual for Systematic Reviews of Interventions.^24^ We used a random-effects model for heterogeneity to address study participants’ different backgrounds and the variety of exercise interventions (which create differences in training variables such as exercise intensity and duration, session frequency, etc.). Outcome measures also differ among studies depending on the available technology to quantify muscle mass and physical performance. Finally, we expected methodological heterogeneity, as exercise training research uses different designs.

We used inverse variance because we analysed continuous data to compare exercise vs control or conventional care groups using mean and standard deviations from post-intervention values for each outcome. When other forms of variability measures such as standard error or confidence intervals (CI) appeared, we calculated the standard deviations using RevMan 5.4.^24^

We did not plan to investigate subgroups of patients, as we expected the participants to be homogeneous (see inclusion criteria); however, considering the heterogeneous nature of exercise training protocols, we expected to find different types of exercise training, which are classified as either endurance or resistance. In the case of this variety, we planned to perform a subgroup analysis comparing endurance training vs resistance training, investigating heterogeneity across subgroup results and analysing the variability in effect estimates between types of exercise using an I² statistic.

For each outcome analysed, we present the data as mean differences (MD) with a 95% CI. We used standardised mean differences (SMD) to represent effect sizes and facilitate interpretations,^25^ with a value of 0.2 set as small, 0.5 as moderate and 0.8 or higher as a large effect size.^26^ Positive values indicated that it was favourable to the exercise intervention, and vice versa. Zero value suggested that there was no effect on the analysed variable.

### Heterogeneity

We quantified heterogeneity using the χ^2^ test, where a significant *p-value* was indicative of more considerable heterogeneity. We also applied a complementary inconsistency test I^2^ and presented this with its corresponding 95% CI to assess the degree of heterogeneity. Higher I^2^ values indicated greater heterogeneity.^27^

### Risk of Bias

We assessed all the included RCTs using the Cochrane collaboration risk of bias tool to search for elements that could over- or under-estimate the intervention’s effect. We evaluated the following for each study: selection bias (random sequence and generations, allocation concealment), performance bias (blinding of participants and research staff), detection bias (blinding of outcome evaluation), attrition bias (incomplete outcome data), reporting bias (selective reporting), other sources of bias.^24^ Two independent reviewers (AG and OA) performed the assessments; they discussed discrepancies until reaching a consensus.

## RESULTS

### Review studies

We initially identified 66 articles in PubMed, CINAHL and Scopus. Of these, we excluded one by duplication and 29 after analysing the title and abstract. Among the remaining 36 articles, we excluded 29 due to the combination of exercise with another intervention (n = 15), participants under 18 years (n = 2), article written in Spanish (n = 1), reports not RCTs (n = 5), no determination or evaluation of sarcopenia criteria (n = 2) and patients were not diagnosed with NAFLD or had comorbidities (n = 4). Supplementary Material 3 lists the excluded studies and causes for the exclusion.^28–55^ We included seven articles in the final analysis^56–62^ (Figure 1).

**Figure 1.**
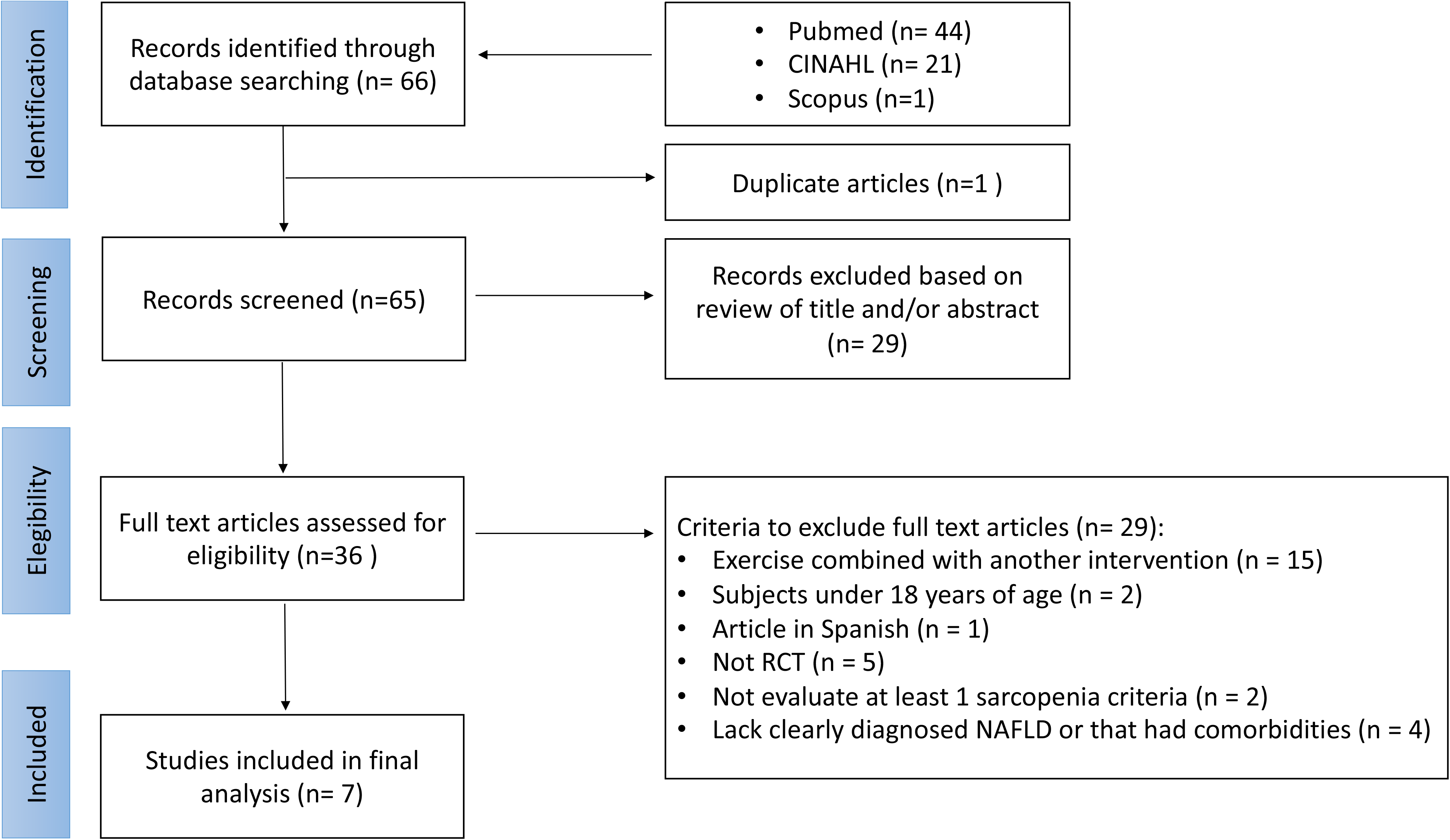
Flow chart of search results and study selection according to the PRISMA statement.

### Description of included studies

A total of 182 participants were included in the analysis. Among them, 98 participated in exercise interventions and 84 in control groups (see Supplementary Material 2). Five studies used a moderate-intensity protocol of endurance training.^56–59 62^ Three studies used a combined exercise protocol (endurance and resistance);^59–61^ In all seven studies the exercise interventions were supervised. All the studies had a control group composed of patients with NAFLD following standard or conventional care, which did not include exercise intervention or any other component that could have influenced the outcomes of interest.^56–62^

Exercise intervention duration ranged from 12–16 weeks, with training frequency of 2–5 times per week lasting 20–60 min per session. Endurance training intensity varied from 45–75% of oxygen consumption peak (VO_2peak_), 30–60% of heart rate reserve (HRR) or 16–18/20 (Borg Scale) rating of perceived exertion (RPE). Endurance training varied: cycle ergometer, treadmill, Nordic brisk walking, walking and self-selected gymnasium exercise routines.

Two articles described high-intensity interval training (HIIT) as a form of endurance training. One of these articles used 5 intervals of 2 min, adding 10 s to each range per week with 3 min of recovery and an RPE (Borg Scale) intensity of 16–17 (very hard).^60^ The other article used cycling intervals with an RPE (Borg scale) intensity of 16–18.^61^ The remaining five studies carried out continuous training.^56–59 62^

Among the three articles that carried out combined training, one performed a moderate-intensity endurance protocol (40%–60% HRR) combined with resistance training; however, the details of the latter training were not described.^59^ Another study used combined training protocol by performing endurance interval training with an intensity based on RPE of 16–18 (very hard) together with resistance training with an RPE intensity of 14–16 (hard). The exercises included hip and knee extensions, horizontal rows, chest presses, vertical rows and knee extensions.^61^ One article required participants to perform an interval on a cycle ergometer, followed by a light-band-resisted upper body exercise (60 s) in the following order: face-pull, horizontal push, horizontal pull, 30° push. However, it was unclear whether this protocol was a combined workout.^60^

### Quality assessment analysis

The TESTEX scale analysis yielded a median score of 11 out of 15 possible points for the studies,^10^ with values closer to 15 of higher quality than values closer to 1. We evaluated assessment quality by organising the studies as low-quality if their score was less than the median and high-quality if the score was greater than or equal to the median.^10^ Accordingly, six studies were high quality and one article was low-quality (Table 2). The main weaknesses TESTEX revealed were the supervision of physical activity in the control groups (0 of 7 articles), intention-to-treat analysis (1 of 7 studies) and blinding of the assessor for at least one key outcome (1 of 7 articles).

### Risk of bias assessment

The risk of bias was low in the RCTs included in this systematic review and meta-analyses for every outcome. However, we had some concerns about their analysis of physical performance regarding blinding the assessors. Only one of the studies performed this key methodological feature, indicating a potential risk of detection bias for articles that used a cardiopulmonary exercise test (CPET) to assess physical performance, as assessors might influence patients’ performance in this type of evaluation. The details about the risk of bias assessment can be found in Supplementary Materials 4 and 5.

### Change in muscle strength

We excluded the muscle strength criterion for sarcopenia because none of the selected studies evaluated it.

### Change in physical performance

For the physical performance criterion, we performed a meta-analysis with the results obtained post-exercise by comparing the control group vs the exercise group. Data on physical performance were present in four studies,^56–59^ all of which determined the direct maximum oxygen consumption (VO_2max_) or oxygen consumption peak (VO_2peak_) based on CPET and expressed in mL/Kg*min. In this regard, we excluded one article because it performed the 2 Km walking test, which does not directly evaluate physical performance.^62^

Three studies showed significant increases in VO_2max_ or VO_2peak_.^57–59^ One study showed no change; here, the authors recognised that the results could be explained by a type II statistical error because of the small number of participants.^56^

The pooled analysis of the studies indicated a change in physical performance in favour of exercise (MD 8.26 mL/Kg*min [95% CI 5.27–11.24 mL/Kg*min], p < 0.0001, Figure 2) and a large effect size (SMD 1.10 [0.60–1.60], p < 0.0001, I^2^ = 0%). Studies that analysed physical performance had low heterogeneity (Chi^2^ 2.43, p = 0.49; I^2^ 0%; Tau^2^ = 0.00).

**Figure 2.**
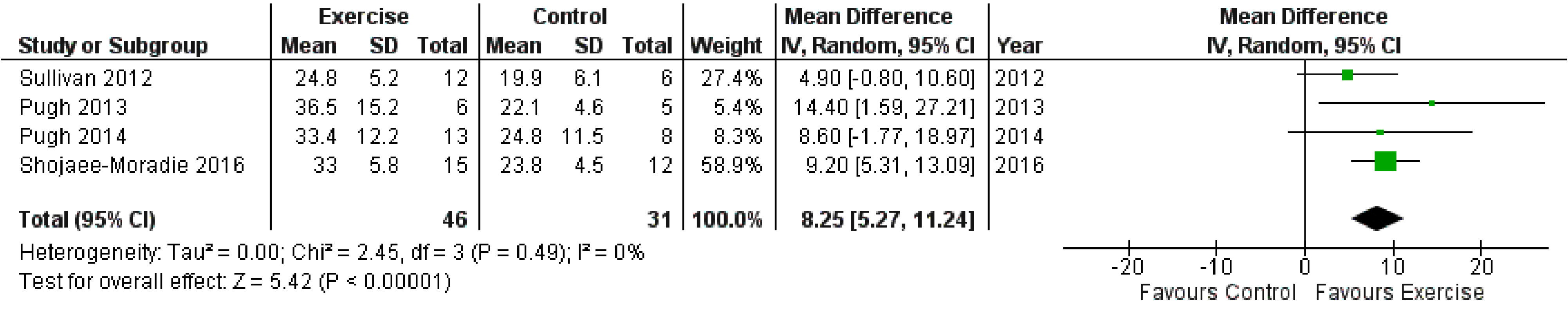
A meta-analysis of pooled effect size and confidence intervals (CIs) (95%) of the interventions with exercise versus control group on physical performance evaluated by the cardiopulmonary exercise test (CPET) (mL/Kg * min).

### Change in muscle mass

Four studies determined muscle mass as LBM.^56 60–62^ They evaluated LBM by different methods, including dual-energy X-ray absorptiometry (DXA) and air displacement plethysmography. Despite this, we could compare the values they reported because they expressed the results in kilograms (Kg). None of these studies reported changes in LBM.

The studies we used to analyse this outcome had low heterogeneity (Chi^2^ 1.74, p = 0.63; I^2^ 0%; Tau^2^ = 0.00); our meta-analysis found no evidence of a difference in the effect between exercise and control groups (MD 1.01 Kg [95% CI –1.78 to 3.80 Kg], p = 0.48, Figure 3). There was a small effect size (SMD 0.09 [−0.27 to 0.44], p = 0.64, I^2^ = 0%).

**Figure 3.**
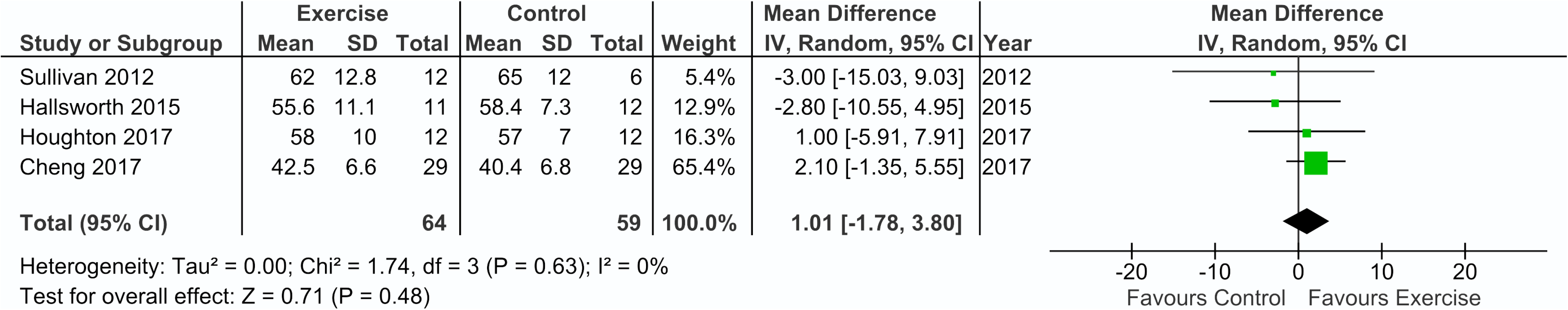
A meta-analysis of pooled effect size (ES) and confidence intervals (CIs) (95%) of the intervention with exercise versus control group on lean mass evaluated by dual-energy X-ray absorptiometry (DXA) and air displacement plethysmograph (Kg).

## DISCUSSION

Following our results, we established the effect of exercise training in two of the three criteria for sarcopenia in NAFLD patients. Exercise training has a positive impact on physical capacity and no effect on the LBM of NAFLD patients. It was not possible to determine the impact of exercise on NAFLD patients’ muscle strength.

### Effect of exercise on muscle strength

Some antecedents show the relationship between NAFLD status and muscle strength.^63^ A recent study suggests that a higher hepatic steatosis index (HIS), a clinical feature of NAFLD patients, is associated with lower muscle strength in both sexes.^63^ This antecedent suggests the relevance of muscle strength on NAFLD. Therefore, it is crucial to establish and analyse the influence of factors that could improve muscle strength in patients with NAFLD. There is consistent evidence that resistance training increases muscle strength.^64–66^ Despite clear antecedents that link increased muscle strength with resistance exercise, none of the RCTs included here featured resistance training as a sole intervention. Three studies used a combined exercise protocol (endurance and resistance).^59–61^ Despite the combined training, we expected them to evaluate physical qualities such as aerobic capacity and muscular strength; however, all the articles only assessed aerobic capacity, and none of them evaluated muscle strength. For this reason, we could not analyse muscle strength as a criterion of sarcopenia.

Beside the conceptual importance of measuring muscle strength in studies that include resistance training, there are also methodological reasons. In general, muscle strength assessments are more straightforward and require less equipment than direct VO_2max_ or VO_2peak_ determination. Concerning the methodology to measure muscle strength, the handgrip strength (HGS) test is one of the most used evidence-based methods for evaluating muscle strength^2^. HGS is a better predictor of adverse clinical outcomes than muscle mass.^67^ If HGS cannot be used, several alternative methodologies are available: knee flexion/extension, dynamometer, one maximum repetition (1RM), ten maximum repetitions (10RM), Isokinetic evaluation and peak expiratory flow (specific to respiration).^2^

One recent study evaluated the relationship between NAFLD status and HGS using a cohort declared as representative of the general Korean population^63 68^. The results indicate that a higher HIS is associated with lower HGS in both sexes.^68^ Therefore, HGS, which is an assessment of general strength in sarcopenia and NAFLD, is quick and easy to perform and should be used as a base procedure for evaluating muscle strength in patients with liver pathologies.^2 68^

Future research should include muscle strength assessments to investigate the effects of exercise, especially resistance training, in the sarcopenia of NAFLD patients.

### Exercise on physical performance

Physical performance, one of the criteria for a sarcopenia diagnosis, is widely evaluated using the CPET, which provides a global assessment of integrative physiological responses.^69 70^ Results from the meta-analysis in our study indicate that most of the articles showed increased physical performance after the training. Only one study found no changes in VO_2peak_ after exercise, which may be explained by the small sample size.^56^

The overall analysis revealed low heterogeneity and an improvement in physical performance under exercise regardless of its features (type, intensity, duration). Three studies performed endurance training without another intervention,^56–58^ while one article performed combined resistance and endurance training.^59^ The endurance training parameters applied in these studies were consistent with the clinical practice guidelines for managing NAFLD from European associations for the study of the liver, diabetes and obesity (EASL, EASD, EASO); they describe a comprehensive lifestyle approach for treating these patients.^71^

All the studies performed CPET and reported the VO_2max_ or VO_2peak_ as indicators of maximal aerobic capacity, which reflects the respiratory and circulatory systems’ ability to supply oxygen to skeletal muscles during exercise.^72^ They predicted an increase in oxygen consumption after endurance training, which induces adaptations in the cardiovascular system and increases mitochondrial biogenesis and capillary density in the skeletal muscles, improving the transport and use of oxygen to generate energy.^64 73^ Zenith et al. in 2014 observed increases in VO_2peak_ in cirrhotic patients after eight weeks of endurance training.^53^ This antecedent is consistent with the fact that the primary adaptation to endurance training is cardiorespiratory fitness improvement.^74^ Therefore, our meta-analysis showed that planned and supervised physical exercise improved physical performance in patients with sarcopenia associated with NAFLD. It is essential to mention that these patients should perform regular exercise to preserve its beneficial effects. This recommendation is based on the substantial benefits exercise has on hepatic metabolism and the fat loss induced by endurance training.^75–77^ Thus, training is a therapeutic strategy for improving cardiorespiratory fitness in fatty liver disease.^78^ Furthermore, regular physical activity before liver transplantation is essential to combatting patients’ immediate stress post-transplant; it is also a critical determinant in long-term health after hepatic transplant.^79^

CPET is the gold standard for assessing cardiorespiratory fitness and functional capacity. Unlike tests that estimate oxygen consumption, CPET measures respiratory, cardiovascular and neuromuscular system function.^69 70^ We excluded one study incorporated in our meta-analysis (in the muscle mass outcome) from the physical performance results, as oxygen consumption was determined from a 2 Km walking test.^62^ In addition to the CPET assessment, the studies on patients with sarcopenia associated with NAFLD could include other validated muscle function tests for primary sarcopenia. For example, the short physical performance battery test (SPPB) evaluates gait, strength and balance through a performance scoring system – it is an easy and quick test to implement.^2 80^ SPPB can assess the severity of sarcopenia as well as predicts mortality on the liver transplant waitlist.^81 82^ Future studies should use the SPPB to gain additional information on muscle function in NAFLD patients, as the European Association for the Study of the Liver (EASL) suggests.^83^

### Effect of exercise on muscle mass

As shown in Figure 3, exercise does not affect NAFLD patients’ LBM – a conclusion based on our analysis of four studies.^56 60–62^ These studies determined LBM using plethysmography and DXA. Both methods are widely used to assess body composition in research. The studies showed considerable variability, but the variability across the studies was very consistent. The lack of significance in the overall effect may be because DXA determinations have low reproducibility, dependent upon the equipment used. The gold standard methods suggested for measuring body composition, especially for muscular mass, are CT and magnetic resonance imaging (MRI).^18^

Considering that all the studies in the meta-analysis mainly comprised endurance training protocols as the central exercise intervention, it is possible that endurance training does not affect NAFLD patients’ LBM. Our results are consistent with the findings of a systematic review in cirrhotic patients trained with endurance protocols, which reported no changes in LBM.^9 84^ Contrary to these results, a study in cirrhotic patients featuring resistance training showed an increase in muscle size.^85^ This observation suggests that the type of exercise training determines the effects on muscle mass, and that resistance training could have a positive impact on muscle mass in NAFLD patients.

### Exercise conditions to regulate sarcopenia

Therapies to revert muscular mass loss in NAFLD patients include pharmacological and nutritional approaches. However, exercise-based treatments should not be excluded because they can improve the patients’ generalised physical condition and life quality.^8^ It is widely established that resistance training is the most promising method to increase muscle strength and mass as well as balance;^86^ further, it can induce muscular hypertrophy, translating to increased muscle mass.^86^ Based on these antecedents, resistance training should be indicated as a therapy, alone or in combination, for sarcopenia in NAFLD patients. It is necessary to consider the conditions of resistance training to use in NAFLD patients. The standard conditions suggest that a high load induces a higher hypertrophic response in skeletal muscle;^87^ however, loads below 30% of 1RPM seem sufficient to trigger a hypertrophic response. The current recommendation for hypertrophic training is an intensity of 40–80% of the individual 1RM, with loads > 60% to increase maximal force and muscular mass.^88^ Thus, the best conditions for resistance training in NAFLD patients require further study.

### Limitations

As mentioned, the main limitations of the RCTs included here were difficulty in blinding participants, therapists and assessors and participants’ non-adherence to the assigned intervention regimes. Blinding participants and researchers are a significant challenge in exercise RCTs, as lifestyle interventions require patients and therapists to be fully aware of the type of intervention to be delivered. Nonetheless, for reducing the risk of bias is crucial to blind the outcome assessors, which must be clearly stated in the RCT report. As for non-adherence, researchers should include an intention to treat analyses in a pre-specified fashion, considering that attrition is a common feature in exercise RCTs.

Another limitation was the lack of supervision for not performing the physical activity in the control group. This monitoring is essential to ensuring a low level of physical activity that does not influence the results. If this variable is not controlled, then the sedentary controls could indeed be individuals who belong to a group that performs an unsupervised and undeclared exercise. These limitations should be considered in future RCTs.

The lack of categorisation of NAFLD severity in the RCT participants is another major limitation of the studies included here. This categorisation is fundamental to determine if the effect of exercise training on sarcopenia depends on the stage of the disease. Future RCTs should include this classification when planning and executing exercise protocols.

The exercise training programs were significantly different among the studies regarding exercise intensity, total duration, session frequency and modality (endurance or combined training). Among these, training modality had special consideration, since resistance training is known to be one of the most potent interventions for increasing muscle size and quality in healthy and ill subjects.^89^ This fact is caused by increased synthesis of myofibrillar and mitochondrial proteins in untrained individuals and by an increase in strength, muscle mass and effort tolerance.^64–66 74 90^ Nevertheless, none of the studies used resistance training to treat NAFLD patients. Therefore, we strongly recommend including resistance training as the main component of exercise protocols in future research on sarcopenia in NAFLD patients.

## CONCLUSION

Exercise training is a useful tool to treat sarcopenia in NAFLD patients; it increases physical performance in the form of aerobic capacity. Nevertheless, it does not affect LBM. Future research should include muscle strength assessments and resistance training to evaluate the effects of exercise training on sarcopenia in NAFLD patients.

## WHAT IS ALREADY KNOWN?

- Sarcopenia negatively affects NAFLD progression.
- Previous RCTs and meta-analyses have reported beneficial effects from exercise on liver dysfunction in NAFLD patients.

## WHAT ARE THE NEW FINDINGS?

- There are statistically significant effects of exercise interventions on sarcopenia in NAFLD patients.
- Exercise training has a positive effect on physical performance (aerobic capacity) but does not affect NAFLD patients’ LBM. After the initial screening of the RCTs, we cannot include muscle strength as an outcome because none of these RCTs evaluate it.

## Data Availability

N/A

## Contributors

All authors comply with the ICMJE Recommendations of authorship. A.G., C.C-V., F.S., and O.A. had full access to all of the data in the study and takes responsibility for the integrity of the data and the accuracy of the data analysis, O.A. contributed with the study design, to statistical analysis and interpretation of the results, and C.H-S., F.S. and M.V-B. contributed to the study design and writing of the manuscript. All authors approved the final version to be published.

## Acknowledgment

We acknowledge Orlando Flores Guerrero PhD., Faculty of Health and Behavioural Sciences, The University of Queensland, for his input while drafting the manuscript.

**Supplementary Material 1**. Search strategy in PubMed, CINAHL and Scopus databases.

**Supplementary Material 2**. Details of the studies included in the systematic review and meta-analysis.

**Supplementary Material 3**. Causes for exclusion for each excluded randomised controlled trial.

**Supplementary Material 4**. Methodological quality of the included studies in physical performance analysis. Methodological quality of the randomised controlled trials (n = 4) was assessed using the Cochrane risk of bias tool (six evaluation-critical methodological components).

**Supplementary Material 5**. Methodological quality of the included studies in lean body mass (LBM) analysis. Methodological quality of the randomised controlled trials (n = 4) was assessed using the Cochrane risk of bias tool (six evaluation-critical methodological components).

